# RAGnosis: Retrieval-Augmented Generation for Enhanced Medical Decision Making

**DOI:** 10.1101/2025.06.11.25329438

**Authors:** Amir Rouhollahi, Ali Homaei, Aanchal Sahu, Rayan Ebnali Harari, Farhad R. Nezami

## Abstract

We present RAGnosis, a fully offline, retrieval-augmented framework for interpreting unstructured clinical text using open-weight large language models. As a proof of concept, we apply RAGnosis to the task of paravalvular leak (PVL) classification from cardiac catheterization reports, a process that typically requires slow, expert-driven interpretation. The system combines local OCR, semantic retrieval, and instruction-tuned LLMs to generate evidence-backed predictions and explanations grounded in real clinical documentation. We evaluate four models (DeepSeek 70B, Gemma 27B, Mistral 7B, LLaMA 3B) across 100 reports, analyzing classification accuracy, explanation quality, and retrieval relevance. Results highlight tradeoffs between fluency and reliability, with DeepSeek demonstrating the most consistent performance. By operating entirely on-prem and supporting modular integration, RAGnosis provides a scalable and interpretable foundation for clinical NLP that delivers not just answers but traceable reasoning.

## I. Introduction

Electronic Health Records (EHRs) are the cornerstone of contemporary digital healthcare systems, capturing a heterogeneous mix of structured data, such as laboratory values and billing codes, and unstructured clinical narratives, including free-text notes, diagnostic reports, and scanned handwritten documents. The inherent complexity and scale of EHR data make it a typical example of big data in medicine, necessitating novel computational approaches to extract meaningful insights efficiently. Despite widespread adoption of EHRs, critical information often remains underutilized due to the limitations of conventional data processing techniques and the lack of semantic interoperability across systems [1,2].

To address these challenges, large language models (LLMs) have garnered increasing interest as scalable tools capable of interpreting and synthesizing vast amounts of clinical text. LLMs, especially those built on transformer architectures, exhibit emergent capabilities in summarization, translation, and domain-specific question answering when trained on large corpora through self-supervision [3,4]. However, general-purpose LLMs often falter in clinical environments due to their limited exposure to medical data during pretraining and a lack of instruction tuning aligned with real-world healthcare tasks [4,5]. These limitations have raised concerns regarding factual accuracy, clinical safety, and the potential for hallucinated content.

Retrieval-Augmented Generation (RAG) frameworks offer a promising solution by combining two discrete steps: semantic retrieval of relevant documents from a trusted corpus and conditioned text generation by an LLM [6,7]. This dual-stage approach significantly enhances the factual consistency, traceability, and clinical contextualization of generated outputs, making RAG particularly suitable for decision support tasks in medicine [7,8]. In clinical applications, RAG systems have demonstrated the ability to ground responses in authoritative sources, thus improving interpretability and reducing the propagation of misleading information [6].

Although useful, there are several limitations why using such technology can be challenging such as (i) preserving patient data and being able to maintain data privacy since most of the high performance LLMs are commercialized and the process will be done on a third party cloud service which is not ideal for our use case; (ii) LLMs are prone to hallucination and not having access to the current literature and patient documents will make the process less reliable. To address these challenges, we present RAGnosis, an offline, end-to-end RAG-based clinical platform that integrates state-of-theart optical character recognition (OCR), local vector databases, and instruction-tuned, open-source high-performance LLMs. This privacy-oriented RAG module currently runs on our local server at Brigham and Women’s Hospital. Upon receiving a natural-language query from a clinician, the system converts the query into a dense semantic embedding, retrieves the top-ranked segments from an indexed corpus of anonymized clinical notes, and generates a synthesized answer supported by in-text citations. This architecture not only maximizes transparency and accuracy but also ensures compliance with data governance protocols by avoiding external data transfer.

## II. Background and Related Work

Multiple efforts have underscored the efficacy of integrating retrieval-based methods into generative AI workflows for medicine. The Almanac system, for example, demonstrated that augmenting LLM outputs with guideline-based retrieval significantly improved factual alignment and reduced hallucinations in clinical scenarios [7]. Similarly, the CLEAR framework employed named entity recognition to refine document retrieval and improve the relevance of generated summaries, thereby enhancing both specificity and precision [8]. These systems laid the foundation for combining NLP pipelines with structured clinical knowledge.

Despite such progress, the medical community has historically played a passive role in the creation and adoption of LLMs, emphasizing the urgent need for clinicians to actively participate in defining the scope, training data, and evaluation metrics of medical LLMs. Without this involvement, generic models may continue to dominate, introducing risks of biased outputs and unverified clinical recommendations [4]. Moreover, studies have shown that tuning models with task-specific instructions can substantially enhance their performance, especially in domains characterized by complex, jargon-rich language such as medicine [5].

Benchmarking initiatives like MIRAGE have further revealed that RAG performance is highly sensitive to retriever configuration, corpus curation, and task formulation, underscoring the importance of rigorous empirical evaluation [6]. These findings highlight the necessity of systematic testing using real-world clinical scenarios to ensure model reliability and reproducibility [9] provide compelling evidence that adapted LLMs can outperform experienced clinicians in text summarization, suggesting that well-tuned models can alleviate documentation burdens without compromising quality.

Recent research has also explored cost-effective strategies for LLM development. For instance, smaller instruction-tuned models such as Stanford’s Alpaca, trained on curated clinical datasets, have achieved competitive performance at a fraction of the computational cost, indicating that scalability and affordability need not be mutually exclusive [4]. Furthermore, RAG-enabled architectures have shown utility in varied tasks, including social media monitoring for pharmacovigilance, clinical trial eligibility screening, and rapid cohort identification, demonstrating their versatility across medical domains [6,8].

RAGnosis builds upon these insights to deliver a clinician-centered, privacy-preserving platform that synthesizes clinical information into actionable, verifiable outputs. By combining iterative retrieval, secure computation, and medical LLMs fine-tuned for local contexts, RAGnosis not only enhances decision making,, but also positions clinicians at the helm of AI deployment in healthcare.

## III. Significance

The significance of this module is multiple folds including but not limited to: (i) clinical significance: by providing relevant and evidence-based institution-specific and patientspecific information supporting the clinical decision-making; privacy and compliance: our RAG module ensures HIPAA compliance and patient data privacy by running the whole pipeline completely offline and locally on our servers at BWH; knowledge integration: the RAG model not only uses the currently available medical knowledge database but it incorporates real patient outcomes and treatment responses. This feedback loop will improve the reliability and relevance of the AI suggestions; (iv) scalability: the module architecture allows seamless integration of new real patient reports as well as newly published medical literature to update the guidelines. For this study, we focus on paravalvular leak (PVL), a high-impact complication that occurs when blood flows between the prosthetic valve and surrounding tissue following surgical or transcatheter aortic valve replacement (TAVR). PVL is common, and while trace or mild cases may be benign, moderate to severe PVL has been associated with hemolysis, heart failure, rehospitalization, and increased long-term mortality [10]. Accurate grading of PVL severity is essential for clinical decision-making but remains a slow, expert-dependent process that requires interpreting complex procedural reports, imaging findings, and hemodynamic data. These observations are often embedded in unstructured text, making large-scale analysis difficult using traditional methods. We have selected PVL classification as the initial focus of the RAGnosis framework, which can be expanded into other use cases over time. We use this framework to automate the interpretation of PVL severity from cardiac catheterization reports. It combines a vector similarity retriever with a locally hosted LLM to identify, explain, and classify PVL from procedural text. Each model receives a patient report, retrieves semantically similar statements from a local corpus, and outputs a PVL classification (e.g., severe, moderate, trace) along with an explanation and textual evidence. We evaluate RAGNosis across four open-weight LLMs DeepSeek 70B, Gemma 27B, Mistral 7, and LLaMA 3.2 on a test set of 100 anonymized reports. Each model is assessed not only for accuracy but also for explanation quality and retrieval relevance. This approach represents a practical step toward interpretable AI in medicine, grounded in real clinical documentation, evaluated across multiple open models, and aligned with privacy-preserving deployment.

## IV. Methods and Experiments

Our pipeline ingests raw cardiology reports (PDF or plain text), converts them to structured text, embeds clinically relevant information, embeds them in a vector store, retrieves the most similar chunks at query time, and finally produces a citation-grounded answer via a locally hosted LLM. The high-level dataflow mirrors the draft schematic in Fig. 1.

**Fig. 1.**
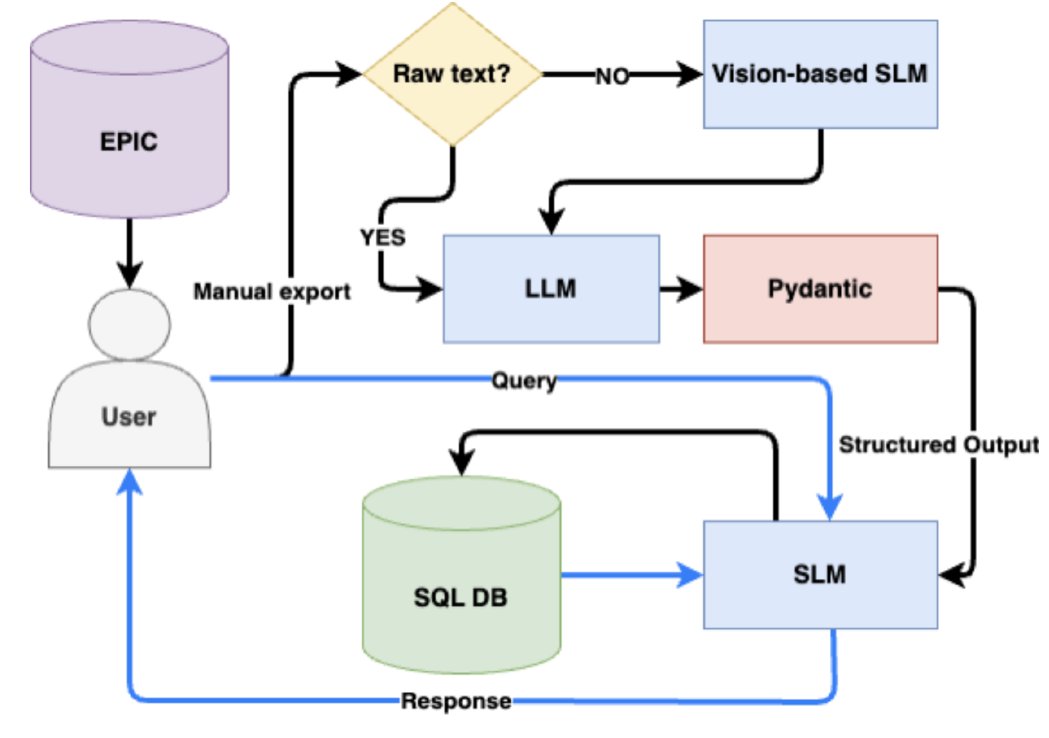
RAGnosis pipeline.

### A. Data

We curated a dataset of anonymized patient catheterization reports (n = 1322) from Brigham and Women’s Hospital. These reports were stored in a local SQLite database and used both for query matching and label verification in the evaluation phase.

Users export reports (such as those from Epic) in PDF or raw text format. When reports are already in raw text format, the content is sent directly to the LLM for analysis. However, when reports are in PDF format, the content must first be processed through an optical character recognition (OCR) module to convert it to raw text. Traditional OCR methods, such as Tesseract OCR [11], face significant challenges when processing images, handwritten documents, low-quality documents, and reports containing equations, tables, or graphs. These conventional approaches lack the necessary accuracy for detecting handwritten content, processing poor-quality documents, and handling complex layouts with tables or noisy backgrounds. To address these limitations, we implemented olmocr vision-based language model [12] to read and convert reports to raw text content. Then, the Python Pydantic library is used to create a consistent structural output in JSON format. The JSON format facilitates easier processing, organization, and integration with our Python codebase, enabling seamless transmission via API to subsequent modules within the RAG-NOSIS package.

### B. Objective

The clinical task was to identify and classify the PVL severity in people who underwent Transcatheter Aortic Valve Replacement (TAVR) surgery, based on unstructured procedural reports as the sole information source. The goal was to evaluate whether LLMs, augmented with retrieval mechanisms, could accurately extract PVL status and generate clinically meaningful explanations. Specifically, the task involved: Identifying and classifying PVL severity from unstructured procedural narratives Generating natural language justifications grounded in relevant clinical text Supporting comparative evaluation of model performance across architectures To perform this task, the LLMs were expected to reason over diverse linguistic markers such as direct PVL labels (e.g., “mild paravalvular leak”), descriptive phrases (“trace paravalvular regurgitation observed near the annulus”), and implicit negations (“no significant regurgitation”), often dispersed across multiple sections of the report. Successful classification required sensitivity to both terminology and contextual cues, including anatomical references, imaging findings, and gradation language that may not explicitly name PVL but imply its presence or absence.

### C. Large Language Models

The four LLMs we used for RAGNosis were:

- DeepSeek 70B (deepseek-r1/70b-32768)
- Gemma 27B (gemma3/27b-32768)
- Mistral 7B
- LLaMA 3.2 (3B)

DeepSeek 70B (deepseek-r1/70b-32768) is a transformer-based, decoder-only language model comprising 70 billion parameters. Released in late 2023 by DeepSeek-VL, the model adopts a GPT-style architecture with rotary position embeddings and multi-head self-attention, and supports a context length of up to 32,768 tokens. This extended window enables the model to process long-form documents without truncating relevant content, a critical requirement for clinical narratives such as procedural reports. Its pre-training was conducted on a large-scale, multi-domain corpus, with subsequent instruction tuning to enhance performance on complex language tasks. It was a strong candidate for this study due to its ability to retain and reason over extended input sequences and its native support for retrieval-augmented generation. Its architectural capacity to integrate retrieved evidence, maintain coherence across long inputs, and produce interpretable outputs aligned well with the demands of clinical document understanding and evidence-linked inference.

Gemma 27B (gemma3/27b-32768) is a 27-billion parameter decoder-only language model released by Google DeepMind in early 2024. Positioned as part of Google’s open-weight model family, Gemma was designed for high-throughput inference with efficient scaling across hardware platforms. The 27B variant supports a context window of 32,768 tokens and incorporates architectural features such as grouped-query attention and activation optimizations to balance performance and resource demands. Pretraining was conducted on a filtered, high-quality dataset with an emphasis on instruction-following and factual grounding. In the context of this study, the model exhibited a tendency toward more verbose responses, often elaborating beyond the immediate evidence. While this occasionally reduced precision in borderline cases, it also contributed to higher scores for readability and linguistic fluency.

Mistral 7B is a lightweight, open-weight language model released in 2023 by Mistral AI. It comprises 7 billion parameters and adopts a decoder-only transformer architecture optimized for efficiency and speed. Notably, it uses sliding window attention and grouped-query attention mechanisms, allowing it to scale effectively across long input sequences without a proportional increase in computational cost. Despite its relatively small size, the model has been shown to match or outperform larger models on several standard language understanding and reasoning benchmarks. In our study, the model produced concise, often highly confident outputs, though its performance was more polarized compared to larger models. It performed well on cases with clear language cues but struggled in more ambiguous or implicit scenarios.

LLaMA 3.2 is a variant within Meta AI’s third-generation LLaMA (Large Language Model Meta AI) family, released in 2024. The model follows a decoder-only transformer architecture and incorporates key improvements over LLaMA 2, including an updated tokenizer, enhanced instruction-following behavior, and support for extended context lengths up to 32,000 tokens. LLaMA 3 models were trained on significantly larger and more diverse corpora, with alignment objectives integrated more directly into the pretraining and fine-tuning process. These updates aimed to improve factual grounding, stability, and usability in open-weight settings. In this study, LLaMA 3.2 (3B parameters) exhibited a mix of high-quality outputs and variable responses, often producing fluent but occasionally less focused explanations. Compared to more heavily instruction-tuned models, LLaMA 3.2’s performance highlighted the impact of prompt sensitivity and grounding strategy when applying general-purpose models to clinical retrieval tasks.

### D. Optimizing Retrieval, Prompting, and Grounding for Clinical NLP

#### Semantic Chunking and Clinical Embedding Optimization

To preserve clinical coherence in long procedural reports, we segment documents into overlapping chunks of 512 tokens with a stride of 128. This design balances retention of semantically relevant information, such as longitudinal mentions of PVL findings, against the constraints of vector store memory and embedding throughput. The overlap helps ensure that contextual cues dispersed across sections (e.g., “moderate leak was later observed…”) are not lost at chunk boundaries, a common failure point in naive token windowing. Each text chunk is transformed into a dense vector using a locally hosted 384-dimensional embedding model optimized for semantic similarity in clinical language. Unlike general-purpose encoders, this model better captures the latent structure of procedural narratives, mapping related expressions such as “trace regurgitation” and “trivial PVL” closer together in embedding space. This improved alignment enhances the retriever’s ability to surface clinically relevant excerpts, especially when synonymous or indirect terminology is used across reports.

#### Prompt Engineering, Grounded Generation, and Explanation Quality

Our system leverages a single-turn prompt template explicitly framed in a clinical role (“You are a cardiologist…”). We found that this instruction, paired with retrieved context, consistently outperformed generic summarization prompts in both explanation quality and faithfulness to evidence. Notably, models without explicit instruction tended to invent pathologies or overstate findings, a phenomenon reduced significantly when role-based grounding was included. Across models, prompt design emerged as a stronger lever than model size alone for optimizing clinical relevance. The generation step is constrained by the top-5 semantically retrieved passages, and each answer is required to cite these excerpts directly, enforcing a soft grounding constraint. We observed that longer explanations did not consistently correlate with higher expert scores. Instead, factual alignment with the retrieved context and accurate citation placement were the strongest indicators of quality. These findings underscore the value of retrieval-augmented generation over freeform outputs, particularly in clinical tasks where subtle variations in language can significantly impact interpretation.

#### Failure Modes and Model Comparison Infrastructure

Qualitative review of model errors revealed that hallucinations clustered in two zones: (1) implied negations (e.g., “no leak was visualized”), and (2) severity gradation mismatches (“mild” vs. “moderate”). To reduce such failures, we implemented fallback logic: if the model response lacks a PVL label or contradicts the retrieved evidence, the system either re-queries with a revised prompt or returns the raw excerpts alone. These mitigations led to a reduction in error rates. By decoupling the retriever and generator modules, RAGnosis supports plug-and-play evaluation across multiple open-weight LLMs. In this study, we benchmarked DeepSeek 70B, Gemma 27B, LLaMA 3.2B, and Mistral 7B using a fixed embedding space and identical context templates. This separation allows us to isolate generative behavior from retrieval quality and supports ablation studies where one module is modified independently, a key step toward standardizing RAG evaluation in clinical NLP.

#### System Modularity and Extensibility

The pipeline is designed with modular isolation across all components - OCR, embedding, retrieval, and generation, allowing each to be independently updated or replaced. This structure enables integration of new models or data formats without requiring full-system retraining and supports deployment flexibility across varied institutional setups. Such extensibility ensures long-term adaptability to evolving clinical standards, data types, and compliance requirements.

## V. Evaluation and Results

The evaluation set comprised 100 explanation and reference pairs generated by four foundation models (DeepSeek = 25, LLaMA = 25, Mistral = 25, Gemma = 25). Each set contained five cases from each PVL category: Not Mentioned, Mentioned Negative, Mild/Trace, Moderate, and Severe. For every case, the model generated a PVL classification, a freetext justification explaining its reasoning, and the exact PDF excerpts used to support its decision. All cases were then shuffled and anonymized before being sent to an MD clinician for a blinded review. The clinician compared each model’s output against the true clinical status and assigned scores from a physician’s standpoint. If a model failed to process a case, it was marked as E (error). If the model’s classification did not match the expert label, it received an F(incorrect). For correct classifications, the reviewer assigned a favorability score from 1 to 4, reflecting how well the model performed. A score of 1 indicates minimal effort or weak justification, and a score of 4 indicates strong, well-supported reasoning. Output from the models was assessed on three criteria:

- Classification Accuracy: Agreement between model prediction and ground truth PVL status
- Explanation Quality: Whether the rationale matched the expert summaries
- Retrieval Relevance: Whether supporting evidence included meaningful clinical statements (qualitative)

Table 1 summarizes the evaluation. LLaMA and Gemma had the highest number of incorrect cases, while Mistral and DeepSeek produced mostly accurate classifications, with DeepSeek demonstrating the most consistent performance across categories.

**TABLE 1.** Evaluation scores for the Llms.

| Scores | E | F | 1 | 2 | 3 | 4 |
| --- | --- | --- | --- | --- | --- | --- |
| DeepSeek | 0 | 2 | 2 | 3 | 18 | 0 |
| Gemma | 1 | 7 | 1 | 3 | 10 | 3 |
| LLaMA | 0 | 7 | 0 | 4 | 9 | 5 |
| Mistral | 2 | 3 | 4 | 0 | 13 | 3 |

## VI. Analysis

### A. Reference Lengths, Explanation Behavior, and Class Balance

The median length of supporting reference excerpts was 267 characters (IQR 198 to 388), while model-generated explanations were typically longer, with a median of 298 characters (IQR 228 to 359). Gemma and LLaMA tended to produce longer explanations than Mistral and DeepSeek across most PVL categories, likely reflecting inherent verbosity differences in their generation styles. However, explanation length showed only a weak to moderate correlation with the length of the source reference (Pearson r: Mistral = 0.34, DeepSeek = 0.26, LLaMA = 0.27, Gemma = 0.10), suggesting that verbosity is not tightly guided by input span. The class balance for model-predicted PVL labels was intentionally uniform; each model produced 5 predictions per label, so performance metrics are not confounded by label prevalence.

### B. Model Accuracy and Expert Favorability

As shown in Fig. 2, DeepSeek achieved the highest overall accuracy (92%), followed by Mistral (87%). LLaMA and Gemma underperformed in comparison, both scoring below 75%. Beyond classification accuracy, expert favorability ratings revealed further distinctions in model behavior. DeepSeek received the most “Good” ratings and no “Poor” scores, indicating strong overall quality and consistency. LLaMA showed the most polarized pattern, earning the highest number of “Excellent” scores but also a significant number of “Fair” ratings. Mistral similarly displayed a broad spread across extremes, while Gemma was rated more narrowly, with most outputs falling into the “Fair” or “Good” categories.

**Fig. 2.**
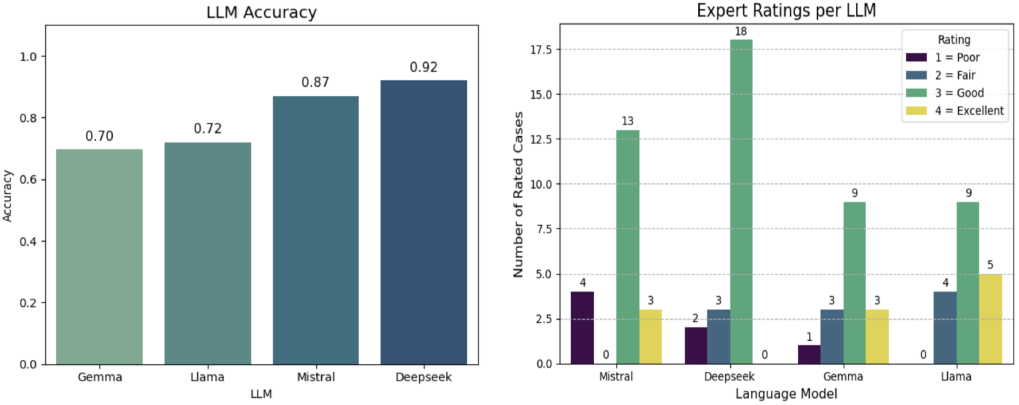
Accuracy achieved by LLMs (left) and Analysis of expert ratings for each LLM (right).

### C. Failure Modes and Error Distribution

Classification errors were concentrated in the “Severe PVL” (40%) and “Not Mentioned” (32%) categories, where models faced challenges with interpreting implicit language around presence or severity. As shown in Fig. 3, no failures occurred in the “Mentioned Negative” category, where denials were explicit. Errors were relatively infrequent in “Mild/Trace” and “Moderate” cases, indicating that misclassifications were most likely at the extremes of the PVL spectrum. Error patterns also varied by model. LLaMA produced the highest number of failures in “Severe PVL” cases, while Gemma struggled most with “Not Mentioned” and “Moderate PVL” examples. DeepSeek had the fewest failures overall, distributed across fewer categories, further emphasizing its robustness in clinical interpretation tasks.

**Fig. 3.**
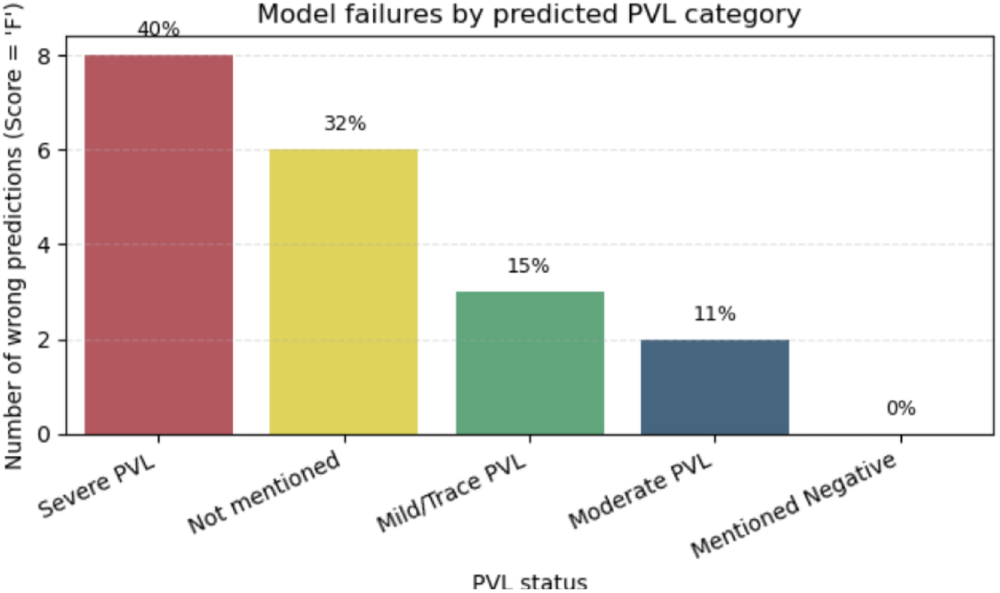
Percentage of wrong predictions by PVL categories.

### D. Expert Ratings and Explanation Quality

Expert ratings varied across models, with DeepSeek showing the most consistent performance. Its scores were tightly clustered around 3 with minimal variance, suggesting steady quality across cases. LLaMA and Mistral, on the other hand, exhibited broader rating distributions and higher medians, indicating less reliability in output quality. Explanation length did not consistently predict expert scores. While top-rated responses had slightly longer median lengths, several short explanations were still rated “Excellent,” and some longer ones received poor scores. This pattern suggests that clarity and clinical relevance, rather than verbosity, were the main drivers of favorable assessments.

### E. Explanation Length Across Models and PVL Categories

Explanation lengths were broadly similar across PVL prediction categories. Median lengths appeared slightly longer for “Mild/Trace” and “Severe PVL” predictions, but with substantial overlap across groups. This suggests that verbosity was not strongly influenced by the predicted class. At the model level, however, there were clear differences, as shown in Fig. 4. Gemma and LLaMA consistently produced longer explanations compared to Mistral and DeepSeek. This trend holds across most PVL categories and likely reflects intrinsic differences in default generation styles rather than task performance. Mistral and DeepSeek, in contrast, generated shorter and more uniform explanations regardless of PVL severity. These differences suggest that some models adjust their verbosity based on content, while others follow a more fixed output strategy.

**Fig. 4.**
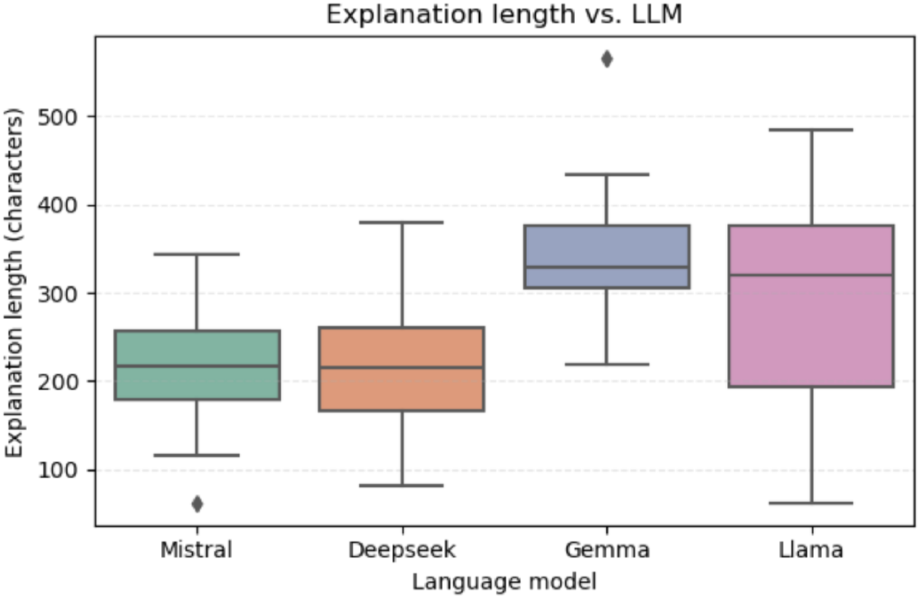
Analysis of explanation lengths of LLMs.

### F. Influence of Reference Length on Generated Justifications

To explore whether explanation verbosity was shaped by the length of supporting evidence, we assessed the correlation between reference and explanation length. As shown in Fig. 5, all models exhibited weak to moderate positive correlations. Pearson r values were 0.34 for Mistral, 0.26 for DeepSeek, 0.27 for LLaMA, and 0.10 for Gemma. While Mistral showed a modest linear trend, the overall findings indicate that reference length was only loosely associated with how much reasoning the model provided.

**Fig. 5.**
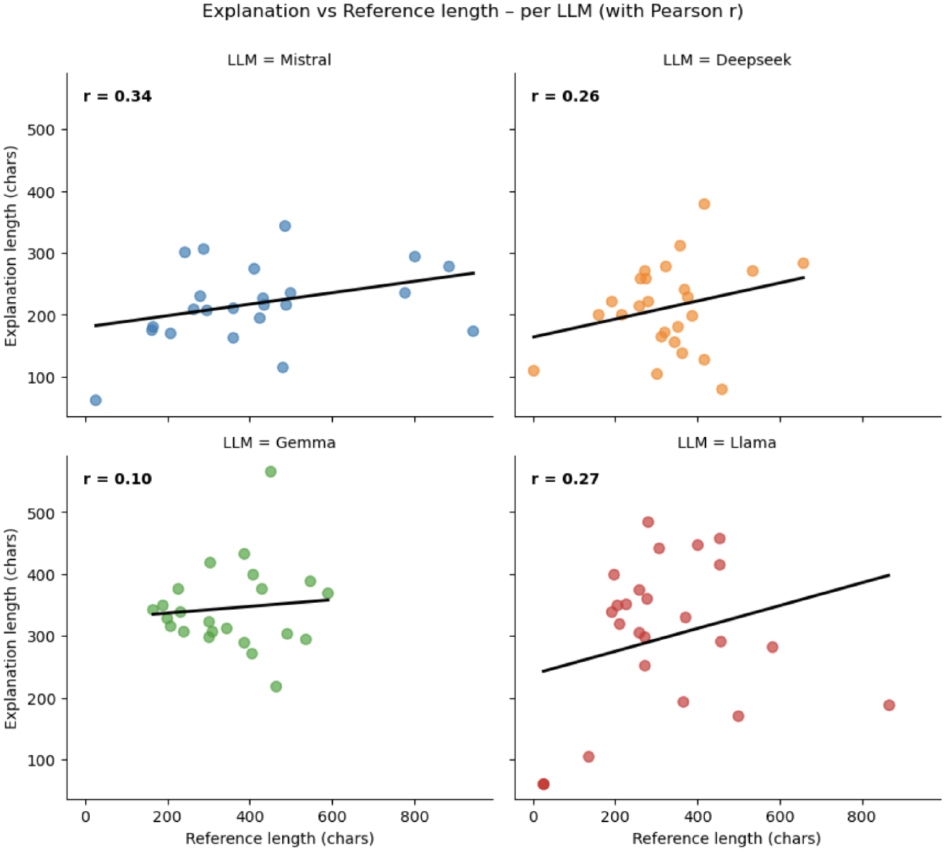
Comparison of reference excerpts from PDF and explanation length.

## VII. Future Work and Limitations

While RAGnosis demonstrates promising performance in medical outcome interpretation and PVL detection, it still has several limitations as a fully local, offline package. First, the training data comes exclusively from the Brigham and Women’s Hospital (BWH) system, so performance on external datasets remains unknown. Clinical documentation practices at BWH may follow institution-specific patterns, which may influence model performance and limit generalizability. Second, the current implementation processes only text; future work will add support for other modalities, such as echocardiography, fluoroscopy, and additional imaging inputs to enrich retrieval and reasoning. Third, real-time inference with large models like DeepSeek requires substantial hardware resources, especially when RAGnosis serves multiple concurrent users. In response, we will upgrade computational hardware and optimize the pipeline so that smaller, more efficient models can deliver comparable performance.

## VIII. Conclusion

We developed RAGnosis as a local, retrieval-augmented platform for extracting and interpreting clinical information from unstructured procedural text. Applied to the task of PVL classification, the system combines semantic retrieval with instruction-tuned large language models to produce evidence-grounded, interpretable outputs. Evaluation across four open-weight models revealed variation in generative behavior, with DeepSeek showing the most consistent performance and Mistral offering strong efficiency–accuracy tradeoffs. By operating entirely offline and supporting modular replacement of key components, the system addresses critical barriers to clinical AI deployment, including privacy, adaptability, and transparency. Although this work centers on PVL, the architecture is readily extensible to other domains where scalable, document-level reasoning is needed. As AI becomes more embedded in care delivery, tools like RAGnosis mark a shift from passive text summarization to active and auditable clinical reasoning, offering not just answers but accountability.

## Data Availability

Due to data privacy, the data used for this research will not be available online.

